# A method for variant agnostic detection of SARS-CoV-2, rapid monitoring of circulating variants, detection of mutations of biological significance, and early detection of emergent variants such as Omicron

**DOI:** 10.1101/2022.01.08.22268865

**Authors:** Eric Lai, David Becker, Pius Brzoska, Tyler Cassens, Jeremy Davis-Turak, Evan Diamond, Manohar Furtado, Manoj Gandhi, Dale Gort, Alexander L. Greninger, Pooneh Hajian, Kathleen Hayashibara, Emily B. Kennedy, Marc Laurent, William Lee, Nicole A. Leonetti, Jean Lozach, James Lu, Jason M. Nguyen, K. M. Clair O’Donovan, Troy Peck, Gail E. Radcliffe, Jimmy M. Ramirez, Pavitra Roychoudhury, Efren Sandoval, Brian Walsh, Marianne Weinell, Cassandra Wesselman, Timothy Wesselman, Simon White, Stephen Williams, David Wong, Yufei Yu, Richard S. Creager

## Abstract

The rapid emergence of new SARS-CoV-2 variants raises a number of public health questions including the capability of diagnostic tests to detect new strains, the efficacy of vaccines, and how to map the geographical distribution of variants to better understand patterns of transmission and possible load on healthcare resources. Next-Generation Sequencing (NGS) is the primary method for detecting and tracing the emergence of new variants, but it is expensive, and it can take weeks before sequence data is available in public repositories. Here, we describe a Polymerase Chain Reaction (PCR)-based genotyping approach that is significantly less expensive, accelerates reporting on SARS-CoV-2 variants, and can be implemented in any testing lab performing PCR.

Specific Single Nucleotide Polymorphisms (SNPs) and indels are identified that have high positive percent agreement (PPA) and negative percent agreement (NPA) compared to NGS for the major genotypes that circulated in 2021. Using a 48-marker panel, testing on 1,128 retrospective samples yielded a PPA and NPA in the 96.3 to 100% and 99.2 to 100% range, respectively, for the top 10 most prevalent lineages. The effect on PPA and NPA of reducing the number of panel markers was also explored.

In addition, with the emergence of Omicron, we also developed an Omicron genotyping panel that distinguishes the Delta and Omicron variants using four (4) highly specific SNPs. Data from testing demonstrates the capability to use the panel to rapidly track the growing prevalence of the Omicron variant in the United States in December 2021.

## Introduction

Since the beginning of the COVID-19 pandemic, variants have emerged that have the potential to evade vaccines, cause diagnostic test performance issues, or cause more severe disease.^1—5^ Monitoring and surveillance of the genetic lineages of SARS-CoV-2 positive samples are critical to the timely identification of emerging variants.^6—8^

SARS-CoV-2 genetic lineages in the United States are primarily monitored by Next-Generation Sequencing (NGS) on a random selection of approximately five percent (5%) of SARS-CoV-2 positive samples.^9,10^ While extremely accurate for the detection of existing circulating variants, NGS does not allow for the timely identification of emerging variants. A more focused approach, such as genotyping using Single Nucleotide Polymorphism (SNP) assays, offers significant advantages in terms of cost, throughput, and efficient results reporting (1 to 2 days turnaround time as compared to 10 to 14 days for NGS). ^11—14^ Additionally, SARS-CoV-2 positive samples with undetermined genotyping variant classification could provide a prescreened sample set for NGS, and potentially allow for early identification of emerging variants.

The National Institutes of Health’s (NIH’s) Rapid Acceleration of Diagnostics (RADx^SM^) initiative created a Variant Task Force (VTF) in January 2021 to assess the impact of emerging SARS-CoV-2 variants on *in vitro* diagnostic testing.^15^ In July 2021, the NIH RADx VTF also initiated an effort to develop a SARS-CoV-2 Polymerase Chain Reaction (PCR) assay for variant agnostic detection of SARS-CoV-2, as well as early detection and monitoring of SARS-CoV-2 variants. The aims of this study were to: 1) identify SARS-CoV-2 markers useful for the detection of SARS-CoV-2 positive samples across all variants; 2) develop a panel of SNP markers that can be used to accurately assign lineages to SARS-CoV-2 positive samples; and 3) implement a genotyping approach for the early detection of new and re-emerging variants that signals when markers need to be updated.

This paper outlines the results of the performance validation of the initial genotyping assay and associated marker sets. Additionally, this paper describes how this approach was rapidly adapted to develop a targeted panel of four (4) mutations—three (3) for Omicron and one (1) for Delta—for the purpose of identifying Omicron, and how this Omicron genotyping panel was implemented in several diagnostic labs. Future results from this testing, as well as the associated statistics and trends, will be made available on a publicly accessible dashboard.^16^

## Materials & Methods

### Marker Selection

Data analysis for identifying SARS-CoV-2 markers was performed using the Variant Analysis for Diagnostic Monitoring (DxM) system (ROSALIND). Genome sequences and metadata used for the selection of markers in this study were obtained through a Direct Connectivity Agreement for complete daily worldwide downloads from the GISAID EpiCov database.^17—19^ Sequences not tagged with the “is_complete’ and sequences with “n_content” of more than 0.05 were excluded. Pairwise whole-genome alignments of all sequences were performed using LASTZ v1.04.02 with NCBI Reference Sequence: NC_045512.2 as the SARS-CoV-2 reference genome.^20,21^ The Bioconductor package for genetic variants, VariantAnnotation v1.20.2, was then used for the translation into amino acids in R v3.3.2, and the identification of amino acid substitutions or frameshifts were used to call a unique mutation incident.^22,23^

Selection of the lineages considered for the marker panel was performed by combining the top 100 most frequent lineages reported worldwide for the 120-day period between May 12, 2021 and September 11, 2021 (data not shown). 1,200,791 sequences representing 393 lineages were analyzed. The top 10 most unique mutations for each World Health Organization (WHO) label were then identified, and multiple combinations of these unique mutations were evaluated to classify a viral sequence into a WHO label with at least 90% overall accuracy. Additional mutations were added to ensure coverage for the Centers for Disease Control and Prevention (CDC) Variants Being Monitored (VBM), Variants of Interest (VOI), Variants of Concern (VOC), and Variants of High Consequence (VOHC) classifications.

The positive percent agreement (PPA) and negative percent agreement (NPA) for each marker set compared to NGS was calculated according to the Clinical and Laboratory Standards Institute (CLSI) EP12-A2: User Protocol for Evaluation of Qualitative Test Performance.^24^ A classifier algorithm was developed to measure the presence, absence, and combination of mutations to accurately assign the WHO label classification. A dedicated system was established to host the classifier algorithm and provide a web application with Application Programmer Interface (API) capabilities for standardized data submission and processing.^25^ This system was established on a secure virtual private cloud instance on the Google Cloud Platform (GCP) with the ability to process thousands of specimens per minute.

### Samples

SARS-CoV-2 positive subject samples used in this investigation were collected in November 2021 and December 2021 by Helix OpCo and The University of Washington (UW), both Clinical Laboratory Improvement Amendments (CLIA)-certified labs participating in the CDC National SARS-CoV-2 Strain Surveillance (NS3) sequencing program to monitor variant distribution in the United States.^26^ Western Institutional Review Board-Copernicus Group (WCG), the Institutional Review Board (IRB) of record for the Helix Respiratory Registry, gave ethical approval for the use of Helix OpCo de-identified remnants of clinical testing. Use of the UW de-identified excess clinical specimens was approved with a consent waiver by the UW IRB.

### Genotyping Assay

A panel of SNP assays was developed using methods previously described.^27^ Primers were selected based on mapping to genome regions with a mutation frequency of less than one percent (1%), ensuring no major polymorphisms interfere with the primers. Primer sets were designed such that amplicon sizes were below 150 base pairs (bp). Minor groove binder (MGB) probes were designed to achieve optimal discrimination between the two (2) alleles by taking the position, nucleotide composition, melting temperature (T_m_), and the type of allele into consideration. The T_m_ of the primers ranged from 59-62°C and the T_m_ of the probes ranged from 59-65°C. Viral RNA was extracted using the MagMAX Viral/Pathogen II Nucleic Acid Isolation Kit (Thermo Fisher Scientific). Real-time reverse transcription PCR using the selected panel was performed using the TaqPath™ 1-Step RT-qPCR Master Mix, CG (Thermo Fisher Scientific) on a QuantStudio™ 7 Real-Time PCR System or ProFlex™ 2 × 384-well PCR System (Thermo Fisher Scientific) followed by endpoint data collection using the QuantStudio™ 7 Real-Time PCR System. Data were analyzed using the TaqMan™ Genotyper v1.6 software (Thermo Fisher Scientific). Normalized reported emission of (Rn) VIC (x-axis) versus Rn FAM (y-axis) from amplification of the reference and mutant alleles was used by the software algorithm to obtain genotype calls. The specific assays for each of the markers are available commercially.^28^

### Next-Generation Sequencing

The SARS-CoV-2 sequencing and consensus sequence generation methods used in this study are previously described.^29,30^

## Results

### Complete Marker Panel

The complete genotyping assay panel consisted of 45 lineage specific markers and three (3) positivity markers (Table 1).

**Table 1.**
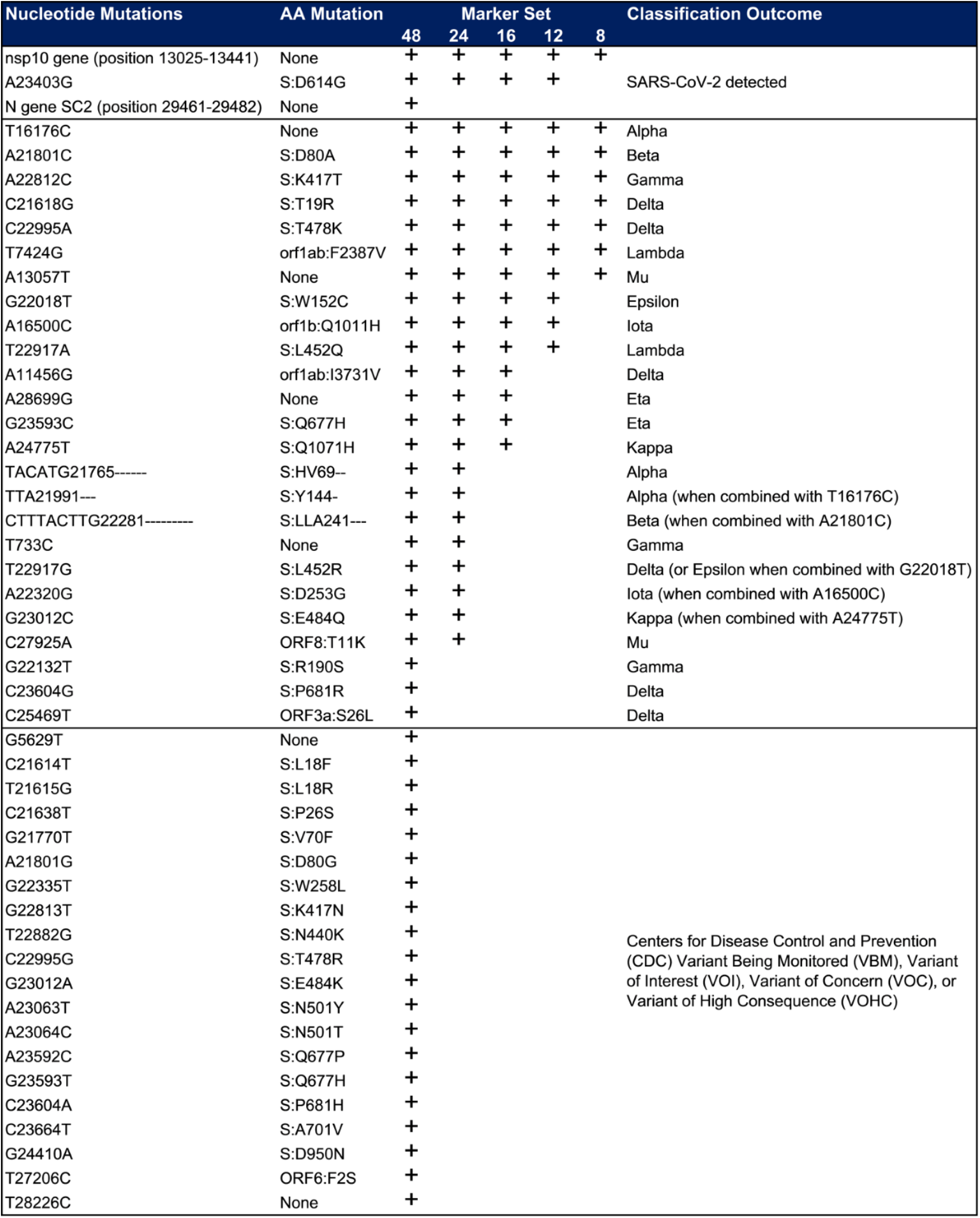
48-, 24-, 16-, 12-, and eight (8)-marker set configurations.

| Nucleotide Mutations | AA Mutation | Marker Set |  |  |  |  | Classification Outcome |
| --- | --- | --- | --- | --- | --- | --- | --- |
|  |  | 48 | 24 | 16 | 12 | 8 |  |
| nsp10 gene (position 13025-13441) | None | + | + | + | + | + | SARS-CoV-2 detected |
| A23403G | S:D614G | + | + | + | + |  |  |
| N gene SC2 (position 29461-29482) | None | + |  |  |  |  |  |
| T16176C | None | + | + | + | + | + | Alpha |
| A21801C | S:D80A | + | + | + | + | + | Beta |
| A22812C | S:K417T | + | + | + | + | + | Gamma |
| C21618G | S:T19R | + | + | + | + | + | Delta |
| C22995A | S:T478K | + | + | + | + | + | Delta |
| T7424G | orf1ab:F2387V | + | + | + | + | + | Lambda |
| A13057T | None | + | + | + | + | + | Mu |
| G22018T | S:W152C | + | + | + | + |  | Epsilon |
| A16500C | orf1b:Q1011H | + | + | + | + |  | Iota |
| T22917A | S:L452Q | + | + | + | + |  | Lambda |
| A11456G | orf1ab:I3731V | + | + | + |  |  | Delta |
| A28699G | None | + | + | + |  |  | Eta |
| G23593C | S:Q677H | + | + | + |  |  | Eta |
| A24775T | S:Q1071H | + | + | + |  |  | Kappa |
| TACATG21765----- | S:HV69-- | + | + |  |  |  | Alpha |
| TTA21991--- | S:Y144- | + | + |  |  |  | Alpha (when combined with T16176C) |
| CTTTACTTG22281----- | S:LLA241--- | + | + |  |  |  | Beta (when combined with A21801C) |
| T733C | None | + | + |  |  |  | Gamma |
| T22917G | S:L452R | + | + |  |  |  | Delta (or Epsilon when combined with G22018T) |
| A22320G | S:D253G | + | + |  |  |  | Iota (when combined with A16500C) |
| G23012C | S:E484Q | + | + |  |  |  | Kappa (when combined with A24775T) |
| C27925A | ORF8:T11K | + | + |  |  |  | Mu |
| G22132T | S:R190S | + |  |  |  |  | Gamma |
| C23604G | S:P681R | + |  |  |  |  | Delta |
| C25469T | ORF3a:S26L | + |  |  |  |  | Delta |
| G5629T | None | + |  |  |  |  | Centers for Disease Control and Prevention (CDC) Variant Being Monitored (VBM), Variant of Interest (VOI), Variant of Concern (VOC), or Variant of High Consequence (VOHC) |
| C21614T | S:L18F | + |  |  |  |  |  |
| T21615G | S:L18R | + |  |  |  |  |  |
| C21638T | S:P26S | + |  |  |  |  |  |
| G21770T | S:V70F | + |  |  |  |  |  |
| A21801G | S:D80G | + |  |  |  |  |  |
| G22335T | S:W258L | + |  |  |  |  |  |
| G22813T | S:K417N | + |  |  |  |  |  |
| T22882G | S:N440K | + |  |  |  |  |  |
| C22995G | S:T478R | + |  |  |  |  |  |
| G23012A | S:E484K | + |  |  |  |  |  |
| A23063T | S:N501Y | + |  |  |  |  |  |
| A23064C | S:N501T | + |  |  |  |  |  |
| A23592C | S:Q677P | + |  |  |  |  |  |
| G23593T | S:Q677H | + |  |  |  |  |  |
| C23604A | S:P681H | + |  |  |  |  |  |
| C23664T | S:A701V | + |  |  |  |  |  |
| G24410A | S:D950N | + |  |  |  |  |  |
| T27206C | ORF6:F2S | + |  |  |  |  |  |
| T28226C | None | + |  |  |  |  |  |

### Variant Agnostic Positivity Markers

The three (3) variant agnostic markers selected to detect SARS-CoV-2 positivity were as follows: 1) the S Gene: D614G (S:A23403G) mutation–a nonsynonymous mutation resulting in the replacement of aspartic acid with glycine at position 614 of the viral spike protein; 2) a conserved sequence in nsp10 (nucleotides 13025-13441); and 3) a conserved sequence identified by the CDC in the N Gene SC2 region (nucleotides 29461-29482).^31^

A total of 1,128 retrospective samples (1,031 SARS-CoV-2 positive and 97 SARS-CoV-2 negative) were evaluated using the variant agnostic positivity markers (Table 2). The combined markers were detected in all but seven (7) of the 1,031 SARS-CoV-2 positive samples. The PPA using any combination of two (2) or more markers is greater than or equal to 98.9% with the criteria being that one (1) marker detected is enough to make a positive call. Additionally, the PPA using one (1) marker is greater than or equal to 96%. There were no false positive results (data not shown).

**Table 2.**
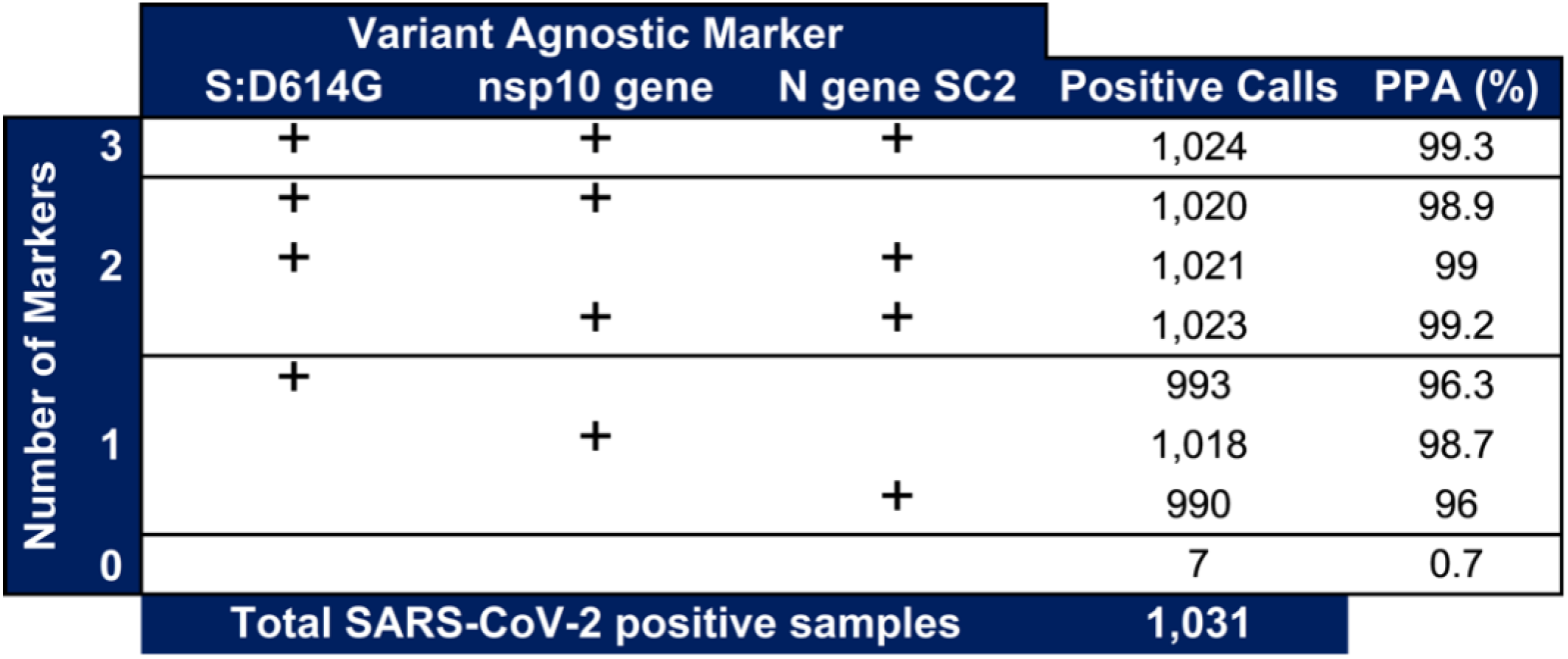
Variant agnostic positivity markers *in vitro* performance.

**Table 3.** 48-marker set *in silico* classifier performance.

|  | 48 Markers |  |
| --- | --- | --- |
|  | Simulated PPA (%) | Simulated NPA (%) |
| Alpha | 99.9 | 98.8 |
| Beta | 97.7 | 100 |
| Gamma | 99 | 100 |
| Delta | 99.8 | 98.1 |
| Epsilon | 96.1 | 100 |
| Eta | 96.2 | 100 |
| Iota | 98.4 | 100 |
| Kappa | 80.7 | 100 |
| Lambda | 98.1 | 100 |
| Mu | 98.8 | 100 |

### Lineage Assignment

The performance of the genotyping assay panel and the associated classifier was determined by *in silico* and *in vitro* studies with retrospectively collected SARS-CoV-2 specimens.

A bioinformatics simulation was performed using GISAID SARS-CoV-2 sequence data from the first week of each month beginning November 2020 through October 2021. 323,148 GISAID sequences were analyzed. With the 48-marker set, simulated PPA ranged from 80.7% to 99.9% and simulated NPA ranged from 98.1% to 100% for the top 10 WHO lineages. The performance for the Kappa variant was impacted by reporting from Asia and Oceania where many Kappa positive samples were misclassified as Delta.

We next determined the clinical performance of the genotyping assay and classifier. The 1,031 SARS-CoV-2 positive samples were genotyped and classified with the 48 markers shown in Table 1. The classifications were then compared to the Phylogenetic Assignment of Named Global Outbreak Lineages (Pango) lineage assignment based on the whole-genome sequences in the GISAID database (Table 4).^32^ The PPA ranged from 96.3% to 100% and the NPA ranged from 99.2% to 100% for the top 10 WHO lineages. The classifier categorized an additional 78 samples as undetermined (data not shown). Pango assigned 77 of these samples to 14 lineages for which the genotyping assay does not include specific markers (Zeta, B.1, B.1.1.507, B.1.2, B.1.221, B.1.241, B.1.517, B.1.596, B.1.609, B.1.625, B.1.628, B.1.634, B.1.637, and C.36.3), and did not classify one (1) of these samples.

**Table 4.** 48-marker set *in vitro* classifier performance.

|  |  |  | Classifier Call |  |  |  |
| --- | --- | --- | --- | --- | --- | --- |
|  |  |  | Positive | Negative |  |  |
| Pango Assignment | Alpha | Positive | 122 | 1 | 99.2 | PPA (%) |
|  |  | Negative | 7 | 908 | 99.2 | NPA (%) |
|  | Beta | Positive | 13 | 0 | 100 | PPA (%) |
|  |  | Negative | 0 | 1,018 | 100 | NPA (%) |
|  | Gamma | Positive | 109 | 0 | 100 | PPA (%) |
|  |  | Negative | 0 | 922 | 100 | NPA (%) |
|  | Delta | Positive | 456 | 5 | 98.9 | PPA (%) |
|  |  | Negative | 1 | 570 | 99.8 | NPA (%) |
|  | Epsilon | Positive | 78 | 3 | 96.3 | PPA (%) |
|  |  | Negative | 3 | 950 | 99.7 | NPA (%) |
|  | Eta | Positive | 27 | 0 | 100 | PPA (%) |
|  |  | Negative | 0 | 1,004 | 100 | NPA (%) |
|  | Iota | Positive | 82 | 0 | 100 | PPA (%) |
|  |  | Negative | 1 | 949 | 99.9 | NPA (%) |
|  | Kappa | Positive | 1 | 0 | 100 | PPA (%) |
|  |  | Negative | 0 | 1,030 | 100 | NPA (%) |
|  | Lambda | Positive | 2 | 0 | 100 | PPA (%) |
|  |  | Negative | 0 | 1,029 | 100 | NPA (%) |
|  | Mu | Positive | 54 | 0 | 100 | PPA (%) |
|  |  | Negative | 0 | 977 | 100 | NPA (%) |

### Marker Reduction

To optimize assay performance in terms of sample input, reductions of the 48-marker panel were explored. We assessed the performance of 24-, 16-, 12-, and eight (8)-marker sets that were defined based on mutation combination performance and targeted lineage prevalence during the 120-day period between May 12, 2021 and September 11, 2021 (Table 5). Each of the panels also included two (2) of the variant agnostic positivity markers (nsp10 gene and S:D614G), which were used as assay internal controls. The 48-, 24-, and 16-marker sets identified the top 10 most prevalent WHO lineages as of September 11, 2021 (Alpha, Beta, Gamma, Delta, Epsilon, Eta, Iota, Kappa, Lambda, and Mu), while the 12- and eight (8)-marker sets identified eight (8) and six (6) of the top 10 WHO lineages, respectively.

**Table 5.**
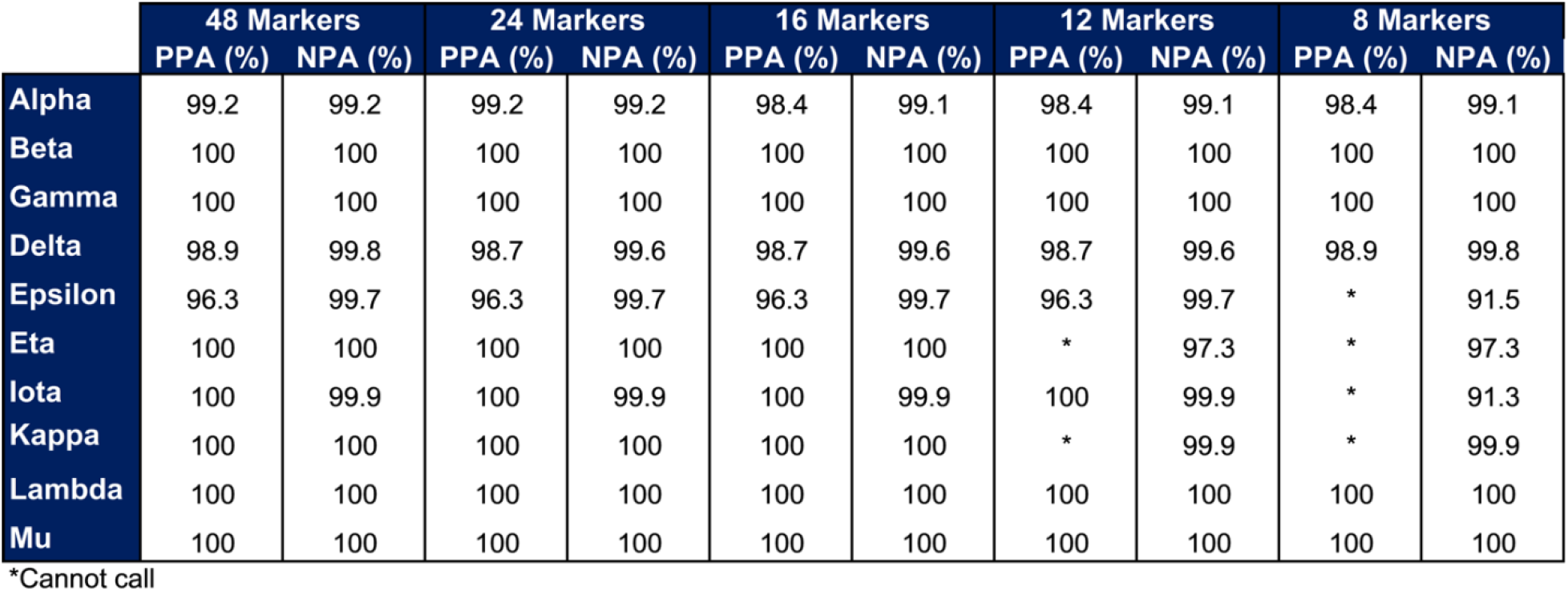
48-, 24-, 16-, 12-, and eight (8)-marker sets *in vitro* classifier performance.

The identification of samples that could not be determined by the classifier was further investigated. The number of undetermined samples for each marker set are shown in Figure 1. The percentage of undetermined samples ranged from approximately seven percent (7%) to 11% for the 12- to 48-marker sets and increases dramatically to 27% with the eight (8)-marker panel.

**Figure 1.**
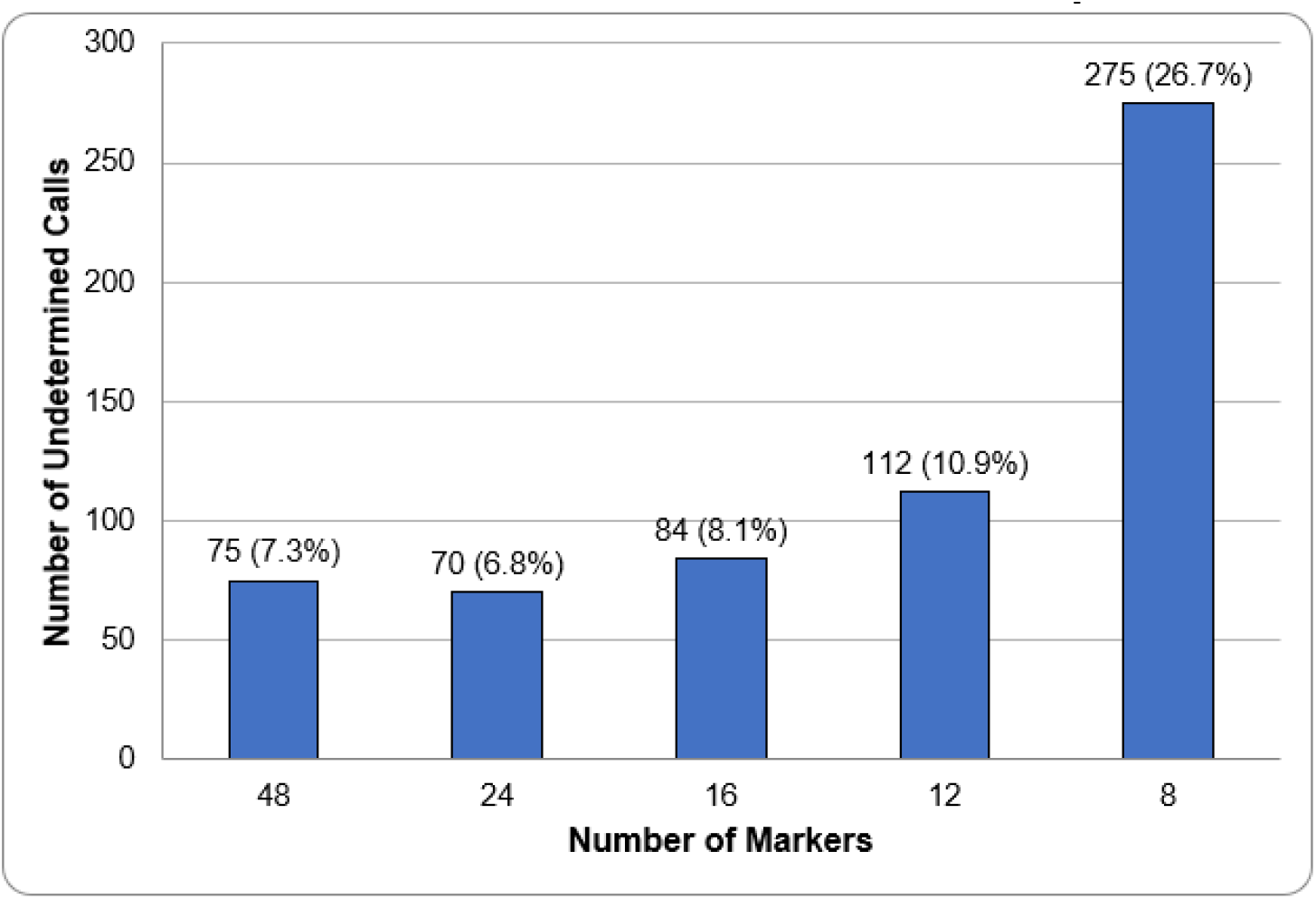
Number of undetermined calls in *in vitro* classifier performance analysis.

### Early detection of new variants

One of the aims of this study was to develop a genotyping approach for the early detection of emerging variants. An increase in the number of undetermined calls by the classifier provides a signal for focused sequencing of those samples, potentially allowing early detection of new variants. To test this hypothesis, a bioinformatics simulation was performed using a modification of the 12-marker panel. The two (2) Delta-specific markers were removed to simulate what would have been observed before and during the emergence of the Delta variant. The number of undetermined calls in the first week of each month from March 2021 through July 2021 are shown in Figure 2. The 10-marker set was able to assign lineages to all positive samples in GISAID for North America in November 2020 and December 2020 (data not shown). The number of undetermined calls was 5, 7, and 1 respectively in January 2021, February 2021, and March 2021. In April 2021, the number increased to 51 followed by a rapid increase over the following three (3) months to 12,825 undetermined calls in July 2021. We then compared these results to the average daily Delta prevalence in the United States from March 2021 to July 2021 as reported by the CDC (Figure 3). The prevalence data for the emerging Delta variant mirrors the rate of increase in undetermined calls over the same period.

**Figure 2.**
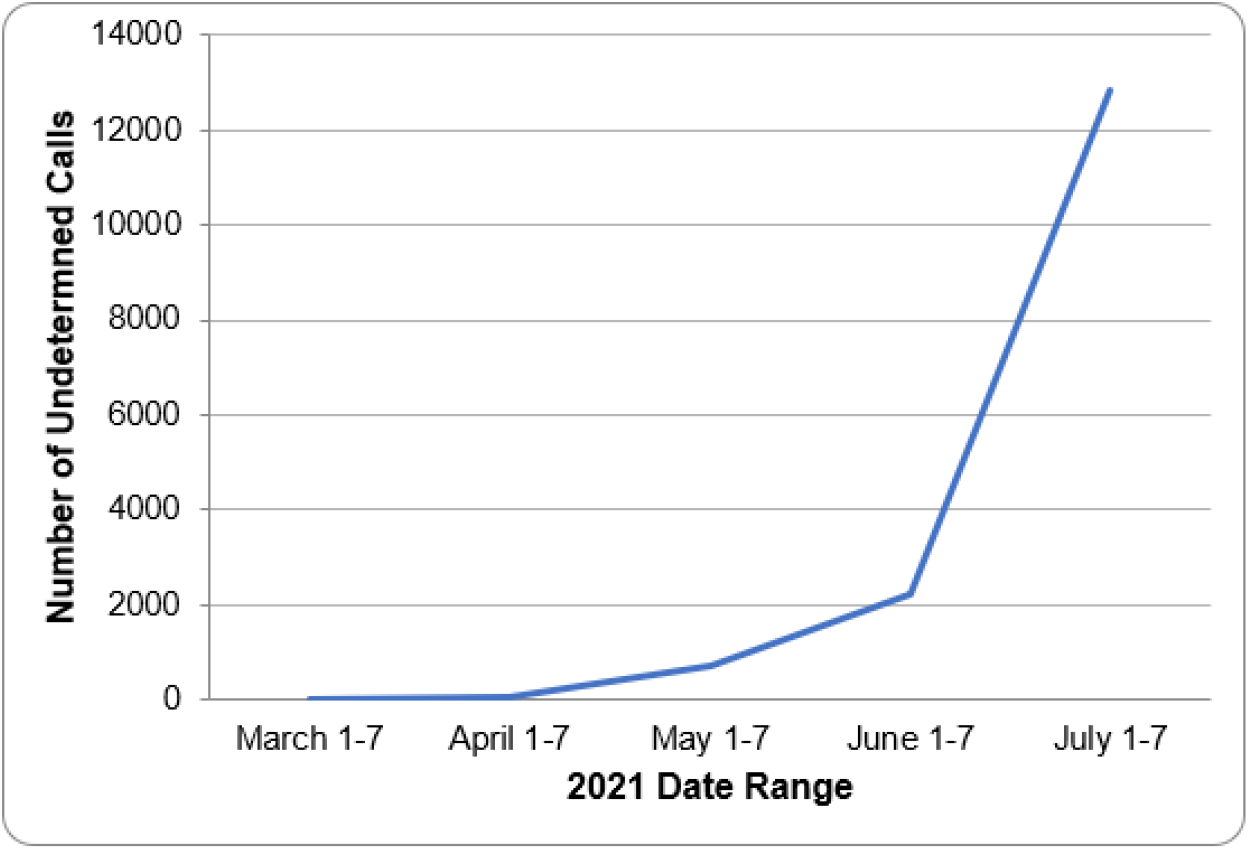
Undetermined calls using 12-marker set without Delta-specific markers.

**Figure 3.**
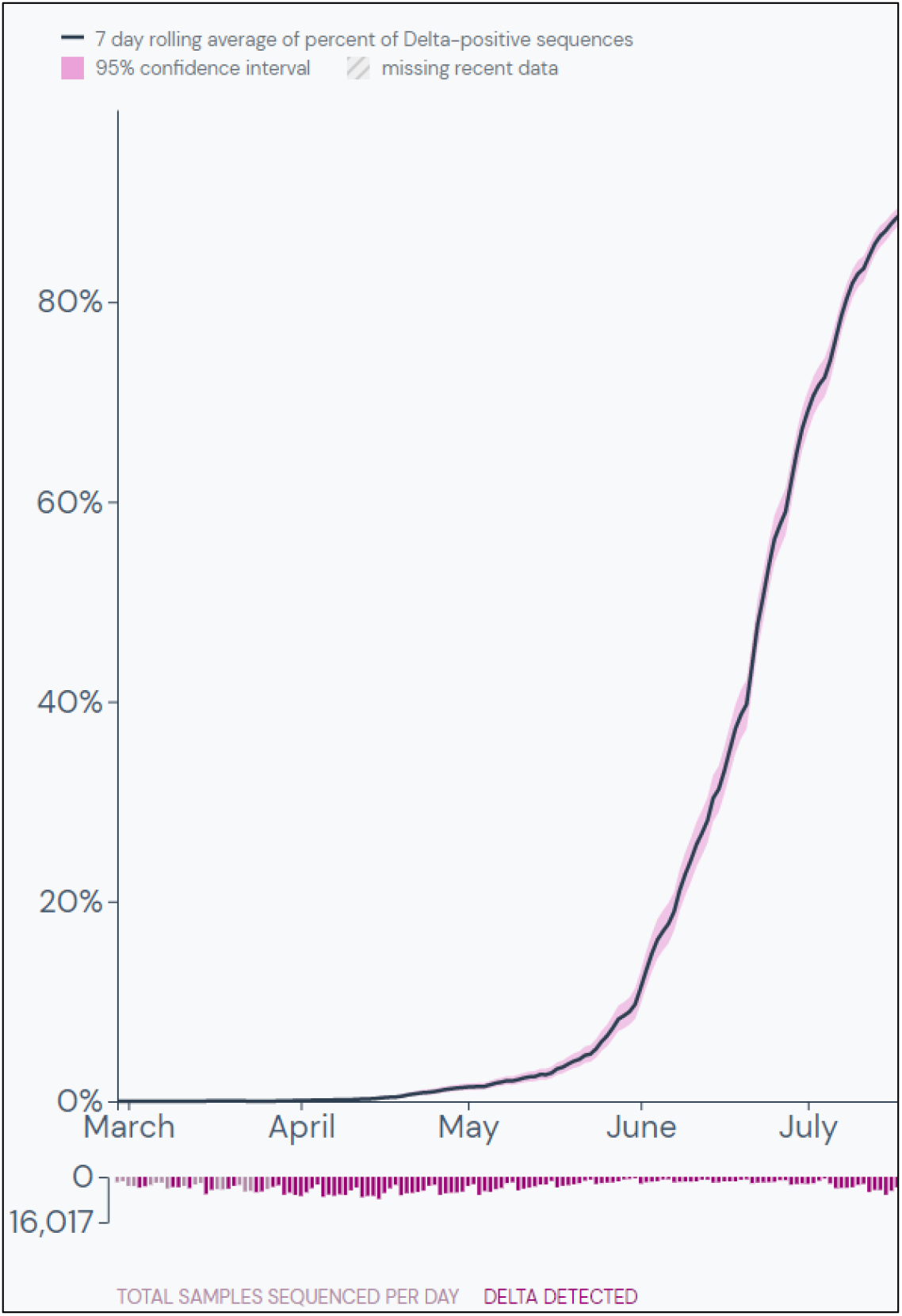
Average daily Delta prevalence in the United States from March 2021 through July 2021. Image courtesy of outbreak.info.^33^.

### Addition of Omicron markers

In response to the emergence of the Omicron variant in November 2021 in South Africa, we rapidly developed markers specific for this newly emerging variant. Sequence analysis of the first 132 Omicron sequences revealed three (3) markers– ORF1ab:A2710T, ORF1ab:T13195C, and S:T547K–found in high percentages of these sequences. Based on *in silico* modeling, there was greater than 99% concurrence between the Pango assignment based on the GISAID sequence and the combined three (3) markers (data not shown). Subsequently, we developed a genotyping assay consisting of the three (3) Omicron-specific markers and one (1) Delta-specific marker (S:T19R).

A total of 1,631 SARS-CoV-2 positive samples were collected and genotyped (Table 6). Sequencing confirmed that these samples consisted of 615 Omicron, 992 Delta, and three (3) B.1 variants, as well as 21 samples that were not classified by Pango. The four (4)-marker panel for Omicron genotyping correctly identified all 615 Omicron samples and 902 of the 992 Delta samples. The 90 Delta samples that were not detected were identified as Delta subtypes by Pango. The four (4)-marker panel classified the three (3) B.1 samples as undetermined, and the 21 samples not classified by Pango as 10 Omicron and 11 Delta.

**Table 6.** Four (4)-marker set *in vitro* classifier performance.

|  |  |  | Classifier Call |  |  |  |
| --- | --- | --- | --- | --- | --- | --- |
|  |  |  | Positive | Negative |  |  |
| Pango Assignment | Omicron | Positive | 615 | 0 | 100 | PPA (%) |
|  |  | Negative | 10 | 1,016 | 99 | NPA (%) |
|  | Delta | Positive | 902 | 90 | 90.9 | PPA (%) |
|  |  | Negative | 11 | 639 | 98.3 | NPA (%) |

### Prevalence of Omicron variant in the United States in December 2021

We next deployed the four (4)-marker panel in two CLIA-certified labs and genotyped 5,372 SARS-CoV-2 positive samples collected mainly in the States of Washington, California, and New York in December 2021. Using the four (4)-marker panel, we determined that the relative prevalence of the Omicron variant grew from approximately 15% on December 9, 2021 to approximately 80% on December 21, 2021, while Delta decreased from 80% to 20% over the same period (Figure 4).

**Figure 4.**
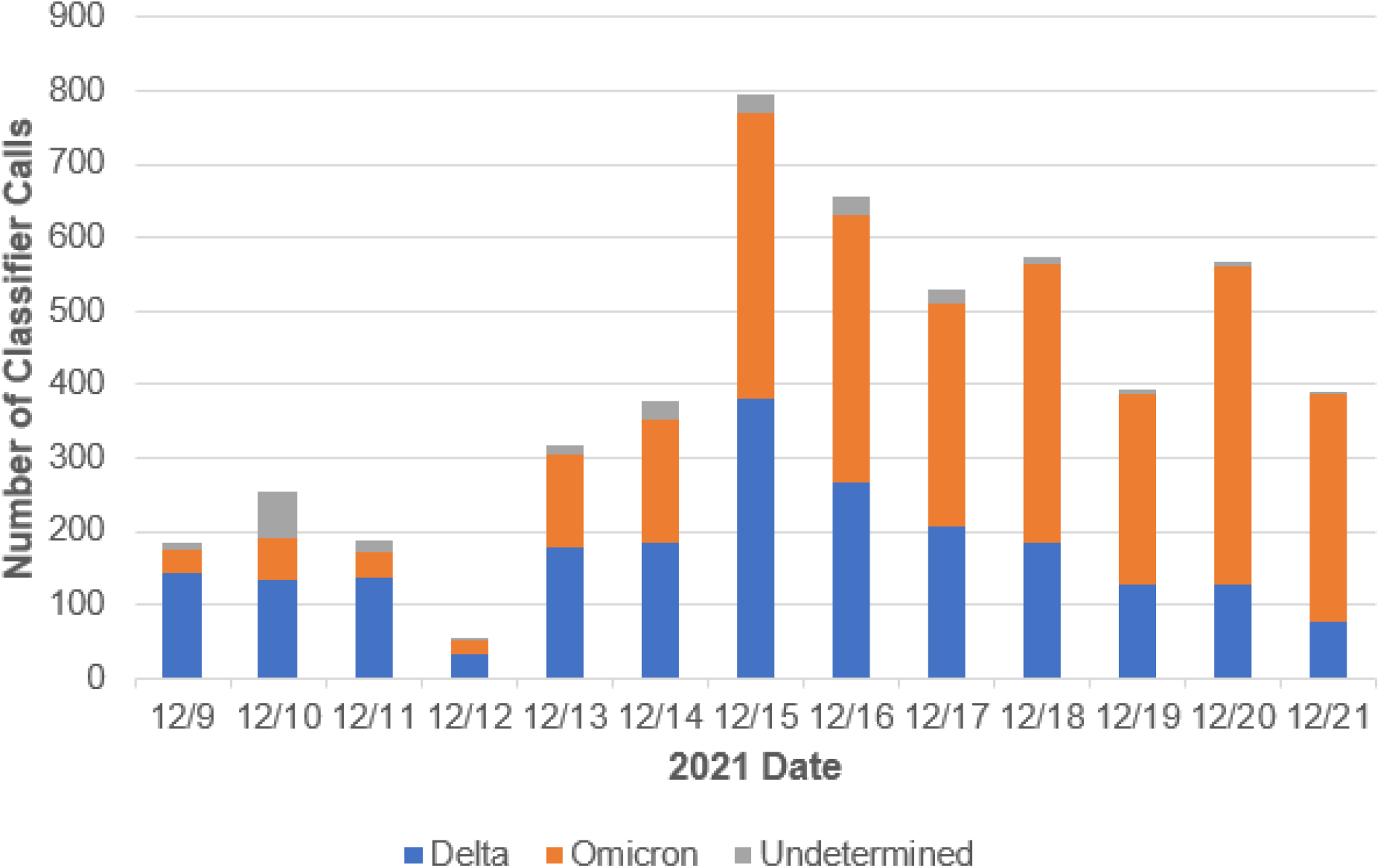
Four (4)-marker set *in vitro* Delta and Omicron classifier calls.

## Discussion

The SARS-CoV-2 virus continues to mutate at an unprecedented scale. Many variants have been reported and several questions are asked each time a new variant emerges. First, can the SARS-CoV-2 diagnostic assays (rapid antigen and/or nucleic acid amplification) detect the new variant? Second, is it possible to develop an experimental approach to monitor the proportion of variants in real time and to detect the emergence of a potential new variant? Finally, are there methods that can be used in conjunction with NGS to provide a faster, cheaper, and more comprehensive view of the prevalence and geographic distribution of variants? In this report, we demonstrated three (3) variant agnostic markers that can detect SARS-CoV-2 positive samples with high PPA and NPA compared to NGS. These markers are present in almost all SARS-CoV-2 samples that were sequenced and should be considered for the development of new assays. We also demonstrated that there are marker combinations that are highly specific for certain variants. Routine use of these genotyping markers could provide early warning that a new or re-emergent variant is circulating. Importantly, genotyping with this assay is a quick and efficient method enabling results reporting in 1 to 2 days compared to 10 to 14 days with NGS. In addition, genotyping assays are less expensive than sequencing. As such, genotyping can be used to monitor a higher percentage of SARS-CoV-2 positive samples than the five percent (5%) random sampling by sequencing currently in practice in the United States. Samples that cannot be assigned to a known variant would be prime candidates for sequencing. Finally, we have demonstrated that the Omicron variant can be identified with high precision with two (2) to three (3) markers. Incorporating the Omicron specific markers with the markers defined to detect previous variants can provide a framework for the detection of the next new variant.

The genotyping markers for Omicron effectively highlighted the transition from Delta to Omicron as the dominant variant. As we illustrated in this report, a static marker set with an omission of the Delta-specific markers experienced a significant decline in accuracy within four (4) months of the emergence of Delta in the United States. To prevent a loss of marker accuracy, we demonstrated an approach to detect new and emerging variants using a classifier algorithm for the recurring analysis of active marker sets across regional and global GISAID sequence data. As emerging variants develop, anomalies in classifier calls and the resulting discordance with sequencing classification will continue to highlight the need for marker modifications. Discrepancies such as these occurred during the retrospective review at the emergence of Delta in the United States. This approach can be applied on a regional or global scale. In this manner, a classifier algorithm would assess the accuracy of current marker sets based on daily analysis of new viral sequences added to GISAID, creating an adaptive and closed-loop process for low-cost, rapid variant identification with emerging variant detection. Indeed, we recently created a free, live dashboard of a real-time genotyping platform.^16^ This illustrates the symbiotic nature of using genotyping markers in conjunction with targeted sequencing. The ultimate goal of these efforts is to provide real-time measurement of variant prevalence with the benefits of increased global awareness and rapid identification of emergent variants.

## Data Availability

All data produced in the present study are available upon reasonable request to the authors.

https://tracker.rosalind.bio

https://www.gisaid.org/

## Acknowledgements

Research reported in this publication was supported by the National Institute of Biomedical Imaging and Bioengineering of the National Institutes of Health (under award numbers 75N92019P00328, U54EB015408, and U54EB027690) as part of the Rapid Acceleration of Diagnostics (RADx^SM^) initiative, launched to speed innovation in the development, commercialization, and implementation of technologies for COVID-19 testing. The funders had no role in the decision to submit the work for publication and the views expressed herein are the authors’ and do not necessarily represent the views of the National Institutes of Health or the United States Department of Health and Human Services.

The authors gratefully acknowledge the originating laboratories responsible for obtaining the specimens, the submitting laboratories where genetic sequence data were generated and shared via the GISAID Initiative, and the GISAID EpiCov Data Curation Team.

Finally, the authors thank Exponent Collaborative for graphic design contributions.

## Notes

### Competing Interest Statement

The authors have declared no competing interest.

### Author Declarations

Helix OpCo samples: Western Institutional Review Board-Copernicus Group (WCG), the Institutional Review Board (IRB) of record for the Helix Respiratory Registry, gave ethical approval for this work. University of Washington Samples: The IRB of the University of Washington (UW) gave ethical approval for this work.

